# Survival After In-Hospital Cardiac Arrest In Critically Ill Patients Implications For The Covid-19 Pandemic?

**DOI:** 10.1101/2020.04.11.20060749

**Authors:** Saket Girotra, Yuanyuan Tang, Paul S. Chan, Brahmajee K. Nallamothu

## Abstract

The coronavirus disease 2019 (COVID-19) outbreak is placing a considerable strain on U.S. healthcare systems. Due to presumptions of poor outcomes in such critically ill patients, many hospitals have started considering a universal do-not-resuscitate order in patients with confirmed Covid-19 given a limited supply of intensive care unit (ICU) beds and the potential risk of transmission of infection to healthcare workers during resuscitation. However, empirical data on survival of cardiac arrest in Covid-19 patients are unavailable at this time.

To inform this debate, we report survival outcomes following cardiopulmonary resuscitation in a cohort of similar critically ill patients with pneumonia or sepsis who were receiving mechanical ventilation in an ICU at the time of arrest. The probability of survival without severe neurological disability (CPC of 1 or 2) ranged from less than 3% to over 22% across key patient subgroups, For patients with an initial rhythm of asystole or PEA, who were also receiving vasopressors at the time of arrest, fewer than 10% were discharged without severe neurological disability (CPC of 1 or 2), and this number dropped to less than 3% in patients over 80 years old. In contrast, survival rates were much higher in younger patients, patients with an initial rhythm of VF or pulseless VT, and in patients receiving ventilatory support without vasopressors.

Our findings suggest caution in universal resuscitation policies. Even in a cohort of critically ill patients on mechanical ventilation, survival outcomes following in-hospital resuscitation were not uniformly poor and varied markedly depending on age, co-morbidities and illness severity. We believe that these data can help inform discussions among patients, providers and hospital leaders regarding resuscitation policies and goals of care in the context of the COVID-19 pandemic.

---

The coronavirus disease 2019 (COVID-19) outbreak is placing a considerable strain on U.S. healthcare systems by requiring both significant acute resources and endangering healthcare team members through airborne infection.^1^ Many hospitals are now considering how to treat COVID-19 patients who suffer cardiac arrest given anecdotal evidence that resuscitation efforts in these individuals may be futile.^2^ However, empiric data on cardiac arrest survival in COVID-19 are not available at the moment. To inform this debate, we report survival data following cardiopulmonary resuscitation in a cohort of critically ill patients with pneumonia or sepsis who were receiving mechanical ventilation in an intensive care unit (ICU) at the time of arrest.

Using Get With The Guidelines-Resuscitation, a U.S. registry of in-hospital cardiac arrest patients,^3^ we identified all adult patients (age 18 years and older) who underwent cardiopulmonary resuscitation for an index in-hospital cardiac arrest. To simulate our study as closely as possible to the COVID-19 population, we restricted our cohort to 5690 patients hospitalized in ICU with a diagnosis of pneumonia or sepsis during the hospitalization and who were receiving mechanical ventilation at the time of arrest during 2014-2018. The study outcomes included survival to discharge, survival with a cerebral performance category (CPC) score of 1 (none to mild neurological disability), and survival with a CPC of 1 or 2 (no worse than moderate disability). We report the above survival outcomes overall, and stratified by patient age (categorized as <50, 50-59, 60-69, 70-79, and ≥ 80 years), initial rhythm (asystole or pulseless electrical activity [PEA] vs. ventricular fibrillation [VF] or pulseless ventricular tachycardia [VT]) and whether patients were receiving intravenous vasopressors at the time of arrest (measure of illness severity). All analyses were carried out using SAS. The study was reviewed by Saint Luke’s Hospital’s Mid America Heart Institute Institutional Review Board which waived the requirement for informed consent.

The median age was 65 years. The initial cardiac arrest rhythm was asystole or PEA in the majority (87%) of patients and more than half (57%) were also receiving intravenous vasopressors at the time of arrest. The overall rate of survival to discharge was 12.5%. Rate of survival with CPC of 1 or 2 was 9.2% and survival with CPC of 1 was 6.2%.

Table includes rates of overall survival, survival with a CPC of 1 or 2, and survival with a CPC of 1 across categories of age-group, initial rhythm and need for vasopressors. Older age, initial rhythm of asystole or PEA and use of vasopressors were associated with lower survival. In patients ≥80 years old with asystole or PEA on mechanical ventilation, the overall rate of survival was 6%, whereas survival with CPC of 1 or 2 was 3.7% and survival with CPC of 1 was 1.7%. Among all patients with asystole or PEA who were also receiving vasopressors (n=2845, 50% of the cohort), less than 10% of patients were discharged with a CPC of 1 or 2 and less than 7% were discharged with a CPC of 1, across all age groups. The corresponding rates of survival with a CPC of 1 or 2 and CPC of 1 were 2.7% and 1.2%, respectively, in the ≥ 80 years age group with asystole/ PEA and on vasopressors. Similar patterns of survival by age and vasopressor use were noted in patients with VF or pulseless VT, although the overall rates were substantially higher compared to patients with asystole or PEA. In patients <50 years of age, with VF or pulseless VT who were not on vasopressors, overall survival was 26.1%, survival with a CPC of 1 or 2 was 22.0%, and survival with CPC of 1 was 16.5%.

We believe that these data can help inform discussions among patients, providers and hospital leaders regarding resuscitation policies and goals of care in the context of the COVID- 19 pandemic which is posing unprecedented challenges to the U.S. healthcare system. The limited supply of ICU beds, mechanical ventilators and personal protective equipment (PPE) is already placing a tremendous strain on health systems. That notwithstanding, a recent article in the *Washington Post* noted that some hospitals are considering implementing a universal do-not-resuscitation orders in patients with confirmed COVID-19 potentially overriding wishes of patients and their families for resuscitation. Furthermore, a recent discussion in the *BMJ* highlighted similar challenges in how to perform resuscitation effectively under these circumstances.^4^

The overall rates of survival and neurological outcomes in this cohort were low, and the survival probability may be even lower in the setting of COVID-19. However, even within our cohort of selected ICU patients who were critically ill with pneumonia or sepsis, large heterogeneity in survival outcomes was present based on patient, cardiac arrest and treatment variables. The probability of survival without severe neurological disability (CPC of 1 or 2) ranged from less than 3% to over 22% across key patient subgroups, while survival with mild to no disability (CPC of 1) ranged from 1.2% to 16.5% Such large variation in survival rates suggest that a blanket prescription of do-not-resuscitate orders in patients with COVID-19 may be unwarranted. Such a policy also ignores the fact that early experience of the pandemic in the U.S. found a nontrivial proportion of hospitalized COVID-19 patients to be <50 years of age and otherwise healthy.^5^ Cardiac arrest in such patients will likely have a different prognosis. Moreover, while asystole or PEA may be more common rhythms in the event of a cardiac arrest in COVID-19 patients due to the associated hypoxia and respiratory failure, patients may also develop ventricular arrhythmias due to myocarditis, and QTc prolongation (e.g., from hydroxychloroquine) which may be reversible. We believe that absent survival data for resuscitation in COVID-19 patients, clinicians could use data on survival presented here to engage patients and families in meaningful conversations regarding the likelihood of survival in the event of a cardiac arrest.

Of course, our findings should be interpreted carefully. Although we selected our cohort to be as closely representative of COVID-19 patients as possible (i.e., patients with pneumonia or sepsis on ventilatory support in an ICU at the time of arrest), the survival rates reported here may represent a ‘best-case scenario’ since COVID-19 patients who arrest maybe sicker. Moreover, resuscitation care in COVID-19 patients in healthcare settings is likely to be delayed due to the need for donning PPE. Second, data on CPC scores were missing in 25.8% of all survivors which was similar across patient subgroups. Therefore, calculations of neurological outcomes were based on the proportion of survivors with CPC 1 or CPC 1 and 2 among those with documented CPC scores. Finally, it is likely that hospitals participating in Get With The Guidelines-Resuscitation are motivated for improving resuscitation care quality and their experience may not be representative of non-participating hospitals.

In conclusion, we found that in a cohort of critically ill patients with pneumonia or sepsis on mechanical ventilation, survival outcomes following in-hospital resuscitation were not uniformly poor. These data argue against a uniform policy of no resuscitation for all COVID-19 patients with in-hospital cardiac arrest and may help guide discussions between patients, providers and hospital leaders in discussing appropriate use of resuscitation during this pandemic, while we await specific data on survival outcomes following resuscitation in COVID- 19 patients.

**Table.** Rates of Survival to Discharge, Survival with a CPC of 1 or 2, and Survival with a CPC of 1 by Age-Group, Cardiac Arrest Rhythm, and Vasopressor Status.

| AGE GROUP | <u>ASYSTOLE / PEA</u> |  | <u>VF / PULSELESS VT</u> |  |
| --- | --- | --- | --- | --- |
|  | All patients | Patients on vasopressors | All patients | Patients on vasopressor |
| **<50 years** |  |  |  |  |
| N | 980 | 562 | 111 | 67 |
| Survival to discharge | 16.8% | 10.1% | 26.1% | 17.9% |
| Survival with a CPC of 1 or 2 * | 12.9% | 8.3% | 22.0% | 15.9% |
| Survival with a CPC of 1 # | 9.9% | 6.3% | 16.5% | 11.9% |
| **50-59 years** |  |  |  |  |
| N | 945 | 533 | 163 | 103 |
| Survival to discharge | 12.1% | 5.1% | 26.4% | 23.3% |
| Survival with a CPC of 1 or 2 * | 8.9% | 3.4% | 19.6% | 18.6% |
| Survival with a CPC of 1 # | 6.3% | 2.7% | 14.5% | 15.5% |
| **60-69 years** |  |  |  |  |
| N | 1305 | 773 | 198 | 107 |
| Survival to discharge | 11.1% | 5.6% | 20.7% | 15.0% |
| Survival with a CPC of 1 or 2 * | 8.2% | 4.0% | 14.3% | 12.3% |
| Survival with a CPC of 1 # | 4.8% | 2.4% | 9.3% | 6.8% |
| **70-79 years** |  |  |  |  |
| N | 1110 | 645 | 169 | 103 |
| Survival to discharge | 8.6% | 4.8% | 20.1% | 13.6% |
| Survival with a CPC of 1 or 2 * | 6.0% | 3.0% | 14.7% | 12.5% |
| Survival with a CPC of 1 # | 3.6% | 2.1% | 10.1% | 7.9% |
| **≥80 years** |  |  |  |  |
| N | 629 | 332 | 80 | 47 |
| Survival to discharge | 6.0% | 3.9% | 15.0% | 10.6% |
| Survival with a CPC of 1 or 2 * | 3.7% | 2.7% | 6.8% | 6.4% |
| Survival with a CPC of 1 # | 1.7% | 1.2% | 5.5% | 6.4% |

Abbreviations: PEA: pulseless electrical activity; VF: ventricular fibrillation; VT: ventricular tachycardia
\* To address missing data on CPC scores, rates of survival with CPC of 1 or 2 were calculated by determining the proportion of survivors with CPC of 1 or 2 among all survivors with available CPC data and multiplying that proportion by the overall survival rate. Similarly, rates of survival with CPC of 1 were calculated by determining the proportion of survivors with CPC of 1 among all survivors with available CPC data and multiplying that proportion by the overall survival rate. Patients who died were assigned a CPC score of 5.

## Data Availability

The study is based on the Get With The Guidelines Resuscitation registry. These data can be available to other researchers with a request from the American Heart Association

